# Pan-cancer analyses reveal molecular and clinical characteristics of TET family members and suggests that TET3 maybe a potential therapeutic target

**DOI:** 10.1101/2024.01.30.24301984

**Authors:** Chunyan Zhang, Jie Zheng, Jin Liu, Guoqiang Xing, Shupeng Zhang, Hekai Chen, Jian Wang, Zhijiang Shao, Yongyuan Li, Zhongmin Jiang, Yingzi Pan, Xiaozhi Liu, Ping Xu, Wenhan Wu

## Abstract

The ten eleven translation (TET) family genes involve a wide range of biological functions in human cancers. However, few studies have comprehensively analyzed the correlation between TET family members and the molecular phenotypes and clinical characteristics of different cancers. Based on updated public databases and integrated several bioinformatics analysis methods, we evaluated expression level, somatic variation, methylation level, prognostic values of TET family gene and explore the association between expression of TET family genes and pathway activity, TME, stemness score, immune subtype, clinical staging, drug sensitivity in pan-cancer. Molecular biology and cytology experiments were used to validate the potential role of TET3 in tumor progression. Each TET family gene exhibits differentiated expression in at least ten detected tumors. The frequency of single nucleotide variation (SNV) of TETs gene was 91.24%, with most missense mutation type, and the main types of copy number variation (CNV) are heterozygous amplification and deletion. The TET1 gene is highly methylated, while the TET2 and TET3 genes are hypomethylated in most cancers, and that closely related to patient prognosis. Pathway activity analysis shows that the TET family genes are involved in multiple signaling pathways such as cell cycle, apoptosis, DNA damage response, hormone AR, PI3K/AKT, and RTK. In addition, the expression level of TET family genes also affects the clinical stage of tumor patients, increases or inhibits the sensitivity of chemotherapy drugs, and then affects the prognosis of patients by participating in the regulation of tumor microenvironment (TME), cellular stemness potential, and the immune subtype. It is particularly pointed out that TET3 plays a role in promoting cancer progression in various tumors, and silencing TET3 can inhibit tumor malignancy and increase chemotherapy sensitivity. These findings may elucidate the role of TET family genes in cancer progression and provide insights for further research on TET3 as a potential target for pan-cancer.

## Introduction

Since the early 20th century, the impact of environmental pollution on human health has been a public health issue of concern worldwide (***de Groot & Munden, 2012***). Environmental pollution mainly harms human health through two ways: one is acute damage caused by exposure to large doses of pollutants in a short period of time; the other is the chronic harm caused by the long-term and continuous accumulation of low-dose pollutants in the body through the air, water, food chain and other means, such as polycyclic aromatic hydrocarbons, phthalates, endocrine disruptors (***Ames & Gold, 1997; Baines et al., 2021; Cao, 2015; Hiatt & Beyeler, 2020***). Long term exposure to polluted environments can cause DNA mutations, and in severe cases, it can cause canceration, deformities, and other abnormalities in human tissues and organs, thereby affecting human health (***Palma-Lara et al., 2020***). Epidemiological studies show that more than 80% of human tumors are closely related to chemical carcinogen in human daily life and work (***Chameides, 2010***).

Despite the continuous progress of surgical and targeted therapies in recent years, a significant proportion of tumor patients still have advanced and widespread metastasis at the time of diagnosis, leading to poor prognosis (***Golemis et al., 2018***). Therefore, in-depth research on the molecular mechanisms of cancer occurrence is of great significance for tumor prevention, early screening and diagnosis, clinical treatment, improvement of patient prognosis and survival rate (***Alqahtani et al., 2019; Ban, Fock, Aryee, & Kovar, 2021; Rodriguez, Schreiber, & Conrad, 2022***).

Genetic variation and epigenetic changes are essential for the occurrence and progression of tumors (***Talib et al., 2021***). Almost all tumors involve single or tandem mutations of a single or multiple gene (***Kossenas & Constantinou, 2021***). To find precise treatment strategies that can effectively kill tumor cells while minimizing damage to normal tissues from a vast array of small molecule chemical drugs, it is necessary to have a comprehensive understanding of the mechanism of action of responsible genes (***Srivastava & Goodwin, 2020***). The creation and development of modern bioinformatics methods provide a shortcut tool for humans to optimally discover and calculate the occurrence of tumors in existing recognized gene information databases, and provide a guideline for treating tumors (***Gonçalves et al., 2021; Nooter & Stoter, 1996; Zaridze, 2008***).

Recent studies have shown that epigenetic regulation such as DNA methylation and demethylation modification is closely related to the occurrence and development of human cancers (***Klutstein, Nejman, Greenfield, & Cedar, 2016; Kulis & Esteller, 2010; Nishiyama & Nakanishi, 2021***). DNA methylation often occurs on the CpG island of the promoter region, leading to oncogene activation and tumor suppressor gene inactivation (***Kulis & Esteller, 2010; Papanicolau-Sengos & Aldape, 2022***). Abnormal DNA methylation can destroy the apoptosis process of cells, make tumor cells insensitive to growth inhibition signals, and induce unlimited replication of tumor cells (***Y. Pan, Liu, Zhou, Su, & Li, 2018***). Therefore, epigenetic research represented by DNA methylation is of great significance and value for tumor molecular diagnosis, prevention and treatment, and prediction of tumor therapeutic effect and prognosis (***C. Chen et al., 2022; Meng et al., 2015***).

Studies have shown that both hypermethylation and hypomethylation are involved in the process of tumor development. The ten eleven translocation (TET) enzymes oxidize 5-methylcytosines (5mCs) and promote locus-specific reversal of DNA methylation (***Kohli & Zhang, 2013***). TET family genes, including TET1, TET2, and TET3, have multiple important biological functions in mammals (***Joshi, Liu, Breslin, & Zhang, 2022; Kinney & Pradhan, 2013; Zeng & Chen, 2019***). The core of the TET family genes is a α-Ketoglutaric acid (α-KG) and Fe^2+^ dependent dioxygenases, which control DNA demethylation, regulate gene transcription, and interfere with life activities such as embryonic development, as well as the occurrence of various diseases such as tumors (***Ma et al., 2021; Rasmussen & Helin, 2016***). Research has also shown that TET family genes can not only induce active demethylation of DNA, but also prevent the spread of methylation by maintaining a low methylation state of DNA, thus closely related to tumors(***Shekhawat et al., 2021***). It is noteworthy that the TET family genes play a dual role in different tumors, with both carcinogenic and anticancer effects. Although TET family genes play an indispensable role in the progression of numerous tumors, there is currently a rare literature on pan-cancer analysis of TET family genes. Therefore, in the current study, we analyzed the relationship between TET family genes and pan-cancer from the aspects of mRNA level, somatic cell variation, DNA methylation, related pathway activity, survival and prognosis, single tumor immune subtype, tumor microenvironment (TME), stem cell index, drug sensitivity, etc., so as to provide important reference data and constructive suggestions based on a comprehensive understanding of the landscape.

## Results

### Expression and differential analysis of TET family gene in pan carcinoma

First, based on the transcriptome data of 33 tumors in UCSC Xena database, we extracted and analyzed the expression of target gene TET family members (namely, TET1, TET2 and TET3) in pan-cancer. The Boxplot results showed that the expression of TET3 gene was highest in pan-cancer, followed by TET2, and the expression of TET1 gene was lowest (Figure 1A, Table 1).

**Figure 1.**
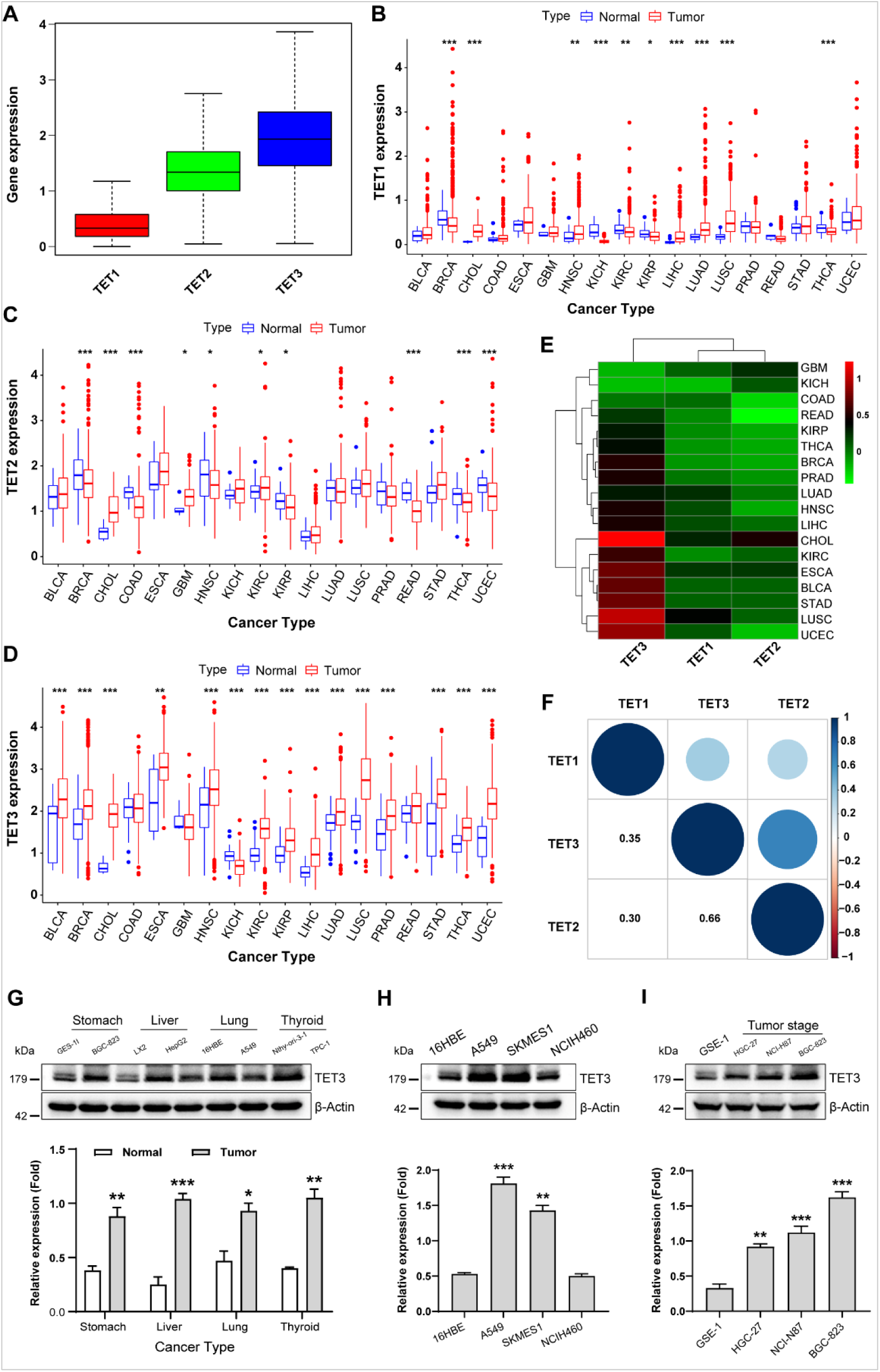
Expression and differential analysis of TET family gene in pan carcinoma. (A) The expression of target gene TET family members (TET1, TET2 and TET3) was analyzed in pan-cancer. **(B-D)** The expression differences of TET1-3 in pan-cancer and its normal tissues. **(E)** The heatmap of the expression differences in each tumor was conducted by Wilcox analysis. **(F)** The expression correlation between TET family genes was analyzed in pan-cancer. **(G-I)** We detected the expression of TET3 protein in different types of cancer cells (G), different pathological subtypes in lung tumors (H) and in different degrees of differentiation of gastric cancer cells (I). Data are presented as the mean ± SD. \**P* < 0.05, \*\**P* < 0.01, \*\*\**P* < 0.001, and ns: no significant.

Next, we screened 18 types of tumors, each containing at least 5 corresponding normal samples, and analyzed the expression differences of TET family members in pan-cancer and its normal tissues. As shown in Figure 1B, TET1 showed abnormal expression in 10 types of tumors, with significantly downregulated expression in BRCA, KICH, KIRC, KIRP, and THCA, and upregulated expression in CHOL, HNSC, LIHC, LUAD, and LUSC tumors than their corresponding normal samples. The expression of TET2, in tumor samples such as BRCA, COAD, HNSC, KIRP, READ, and THCA, is lower, however, in CHOL, GBM, KIRC, and UCEC tumors, is significantly higher than that in corresponding normal control tissues (Fig 1C). The expression of the TET3 gene varies between 15 tumors and corresponding normal samples, including BLCA, BRCA, CHOL, ESCA, HNSC, KIRC, KIRP, LIHC, LUAD, LUSC, PRAD, STAD, THCA, UCEC, and KICH. Among them, except for KICH, TET3 was significantly overexpressed in all 14 other tumors (Fig 1D). It can be seen that the high expression of TET3 may be a key factor in the occurrence, development, and poor prognosis of various clinical tumors.

In order to further clarify the differential expression patterns of TET family members in various tumors, we conducted Wilcox analysis on the expression differences in each tumor and obtained the above differential expression cluster analysis graph (Figure 1E, Table 2). As mentioned earlier, TET3 was significantly upregulated, while TET1 and TET2 were downregulated in most tumors. It is still unclear whether there is a correlation between the expression of three members of the TET family in pan-cancer. Therefore, we conducted a detailed analysis of the correlation between TET family genes in pan-cancer. As shown in Figure 1F, the expression of TET1, TET2, and TET3 is positively correlated in pan-cancer, and the positive correlation between TET2 and TET3 is stronger.

Given the high expression of TET3 in numerous tumors, we speculate that it may be a potential therapeutic target. Therefore, next we detected the expression of TET3 protein in four tumor cell lines (BGC-823, HepG2, A549, TPC-1) and their corresponding normal cell lines (GES-1, LX2, 16HBE, Nthy-ori-3-1). The results showed that the expression levels of TET3 were higher in all four tumor cell lines than the corresponding control cell lines (Fig 1G-I). Then we detected the expression of TET3 protein in three different pathological subtypes of lung cancer cell lines (A549, SKMES1, NCIH460). The results showed that the expression level of TET3 protein in lung adenocarcinoma A549 and lung squamous cell carcinoma SKMES1 was significantly higher than that in normal control and large cell lung cancer cell lines. This result suggests that there is differential expression of TET3 protein in the same type of tumors with different pathological subtypes. Finally, we detected the expression of TET3 protein in gastric cancer cell lines (NCI-N87, BGC-823, HGC-27) with different degrees of differentiation. The results showed that as the degree of cell differentiation decreased, the expression of TET3 protein gradually increased. This suggests that the expression intensity of TET3 protein may indicate the malignant level of gastric cancer.

### Somatic cell variation scenery of TET family genes

To understand the genomic changes of TET1-3 in tumors, we analyzed single nucleotide variation data (SNV, Table 3) and copy number variation data (CNV, Table 4) from a total of 10289 pan-cancer samples from 33 types of tumors, and depicted SNV and CNV landscapes.

Firstly, we analyzed the SNP data related to the TET gene to detect the frequency and variant types of each cancer subtype. As shown in Figures 2A and 2B, among these cancers, the SNV frequency of UCEC, SKCM, and COAD is 19-57%. The SNV frequency of the TETs gene is 91.24% (1052/1153). Variant analysis showed that missense mutation was the main SNP type. SNV percentage analysis showed that the mutation percentages of TET1-3 were 23%, 17%, and 16%, respectively.

**Figure 2.**
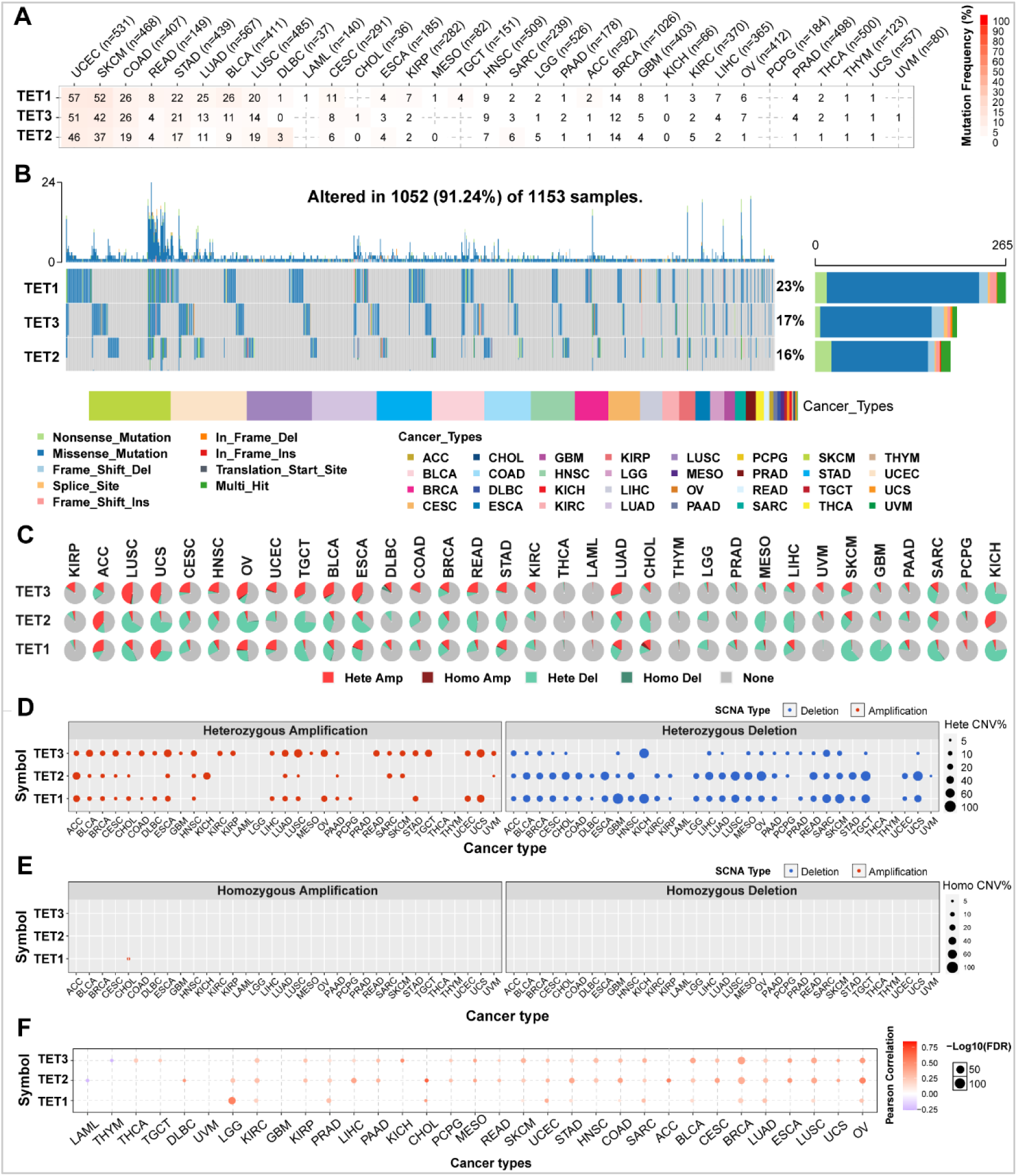
Somatic cell variation scenery of TET family genes. (A) Single nucleotide variation (SNV) frequency of TETs. Numbers in the table represent the number of samples that have the corresponding mutated gene for a given cancer. ‘0’ indicates that there was no mutation in the gene coding region, and no number indicates there was no mutation in any region of the gene. (B) SNV variant types of TETs, which showing the mutation distribution of TETs and a classification of SNV types. (C) Copy number variation (CNV) distribution in 33 cancers. CNV pie chart showing the combined heterozygous/homozygous CNV of each gene in each cancer. A pie chart representing the proportion of different types of CNV of one gene in one cancer, and different colors represent different types of CNV. Hete Amp = heterozygous amplification; Hete Del = heterozygous deletion; Homo Amp = homozygous amplification; Homo Del = homozygous deletion; None = no CNV. (D-E) Heterozygous and Homozygous CNV profile showing the percentage of heterozygous CNVs, including the percentage of amplification and deletion of heterozygous and homozygous CNVs for each gene in each cancer. Only genes with > 5% CNV in a given cancer are shown as a point on the figure. (F) CNV correlation with mRNA expression. The association between paired mRNA expression and CNV percentage in samples was based on a Person’s correlation. The size of the point represents the statistical significance, where the bigger the dot size, the higher the statistical significance. FDR, false discovery rate.

In order to clarify the mutation pattern of CNV, we analyzed the CNV data of TETs genes in the TCGA database. The distribution of CNV pie charts shows that the main types are heterozygous amplification and heterozygous deletion (Figure 2C).

CNV percentage analysis showed that the heterozygous amplification rates of TET1 in ACC and UCS, TET2 in ACC and KICH, and TET3 in ESCA, LUSC, OV, TGCT, and UCS were all greater than 20% (Figure 2D). However, homozygous analysis showed weak homozygous amplification of TET1 only in CHOL (Figure 2E). The heterozygous deletions of TET1 in GBM, KICH, SARC, SKCM, and TGCT, TET2 in CHOL, ESCA, LIHC, LUSC, MESO, OV, READ, TGCT, and UCS, and TET3 in KICH and SARC were all greater than 40% (Figure 2D). No homozygous deletions were observed in all tumors (Figure 2E).

Correlation analysis shows that the mRNA expression of TETs is positively correlated with CNV, especially with TET2 in CHOL and ACC. However, TET2 in LAML and TET3 in THMY are both negatively correlated with CNV (Figure 2F, Table 5). This indicates that CNVs of the TETs genes mediate abnormal expression in pan-cancer, which may play an important role in the progression of cancer.

### Analysis of methylation difference of TET family gene in pan-cancer

We explored the methylation levels of the TET gene in pan-cancer to determine epigenetic regulation. The results showed that there is heterogeneity in the methylation levels of TET family genes in different tumors. The TET1 gene is hypermethylated in most cancers and only hypomethylated in LIHC; The TET2 and TET3 genes exhibit hypomethylation in most tumors. However, TET2 was hypermethylated in THCA, COAD, and KIRP (Figure 3A). The correlation analysis between methylation and mRNA expression showed that the expression of the three genes in pan-cancers was negatively correlated with their methylation levels (Figure 3B, Table 3). Survival analysis showed that methylation of TET1 was associated with low survival rate of ACC and high survival rate of UVM; Methylation of TET2 is associated with high survival rates in ACC, KIRP, and PCPG; The methylation of TET3 is associated with low survival rates in SARC and BLCA (Figure 3C, Table 6). It can be seen that the methylation level of TETs genes is only related to the poor survival rate of a small number of tumors.

**Figure 3.**
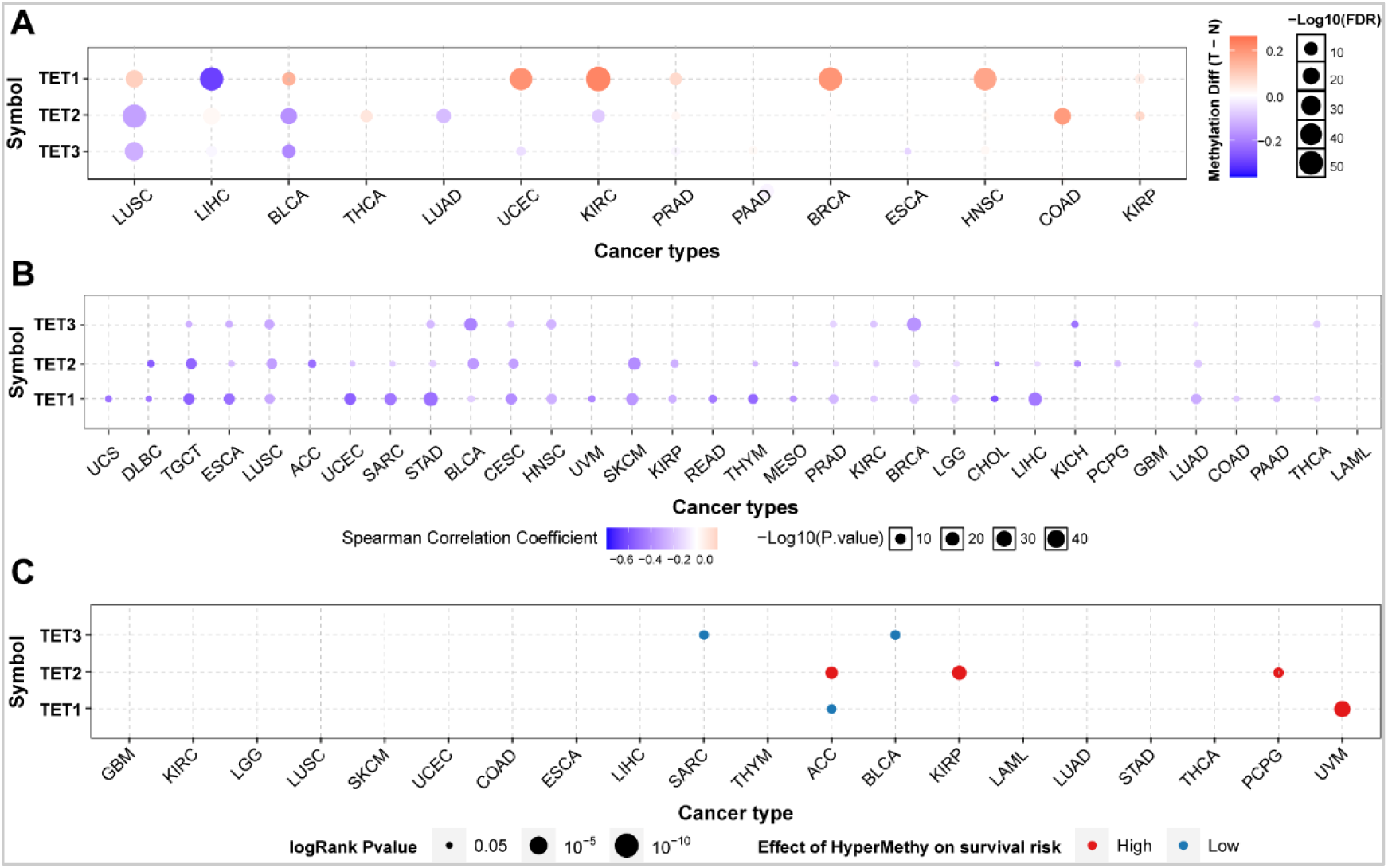
Analysis of methylation difference of TET family gene in pan-cancer. (A) The methylation levels of the TETs genes were explored in pan-cancer to determine epigenetic regulation. (B) The correlation between methylation and mRNA expression of the TETs genes in pan-cancers were further analyzed. Then, we detected the association of survival rate with the methylation level of TETs genes (C).

### Pathway activity analysis of TET family gene in pan-cancer

The analysis of pathway activity showed that TETs were significantly involved in cancer related signaling pathways, including Cell Cycle, Apoptosis, DNA Damage Response, Hormone AR, PI3K/AKT, and RTK. Among them, in addition to participating in cell cycle (16% activation vs. 3% inhibition), apoptosis (16% activation vs. 15% inhibition), and hormone AR (13% activation vs. 6% inhibition), the main activation pathway of TET1 is DNA damage response (19% activation vs. 0% inhibition). TET2 mainly participates in cell cycle (4% activation vs. 28% inhibition) and DNA damage response (10% activation vs. 9% inhibition). TET3 mainly participates in the activation of PI3K/AKT (19%) and the RTK pathway (16% activation vs. 6% inhibition) (Figure 4A and Table 7).

**Figure 4.**
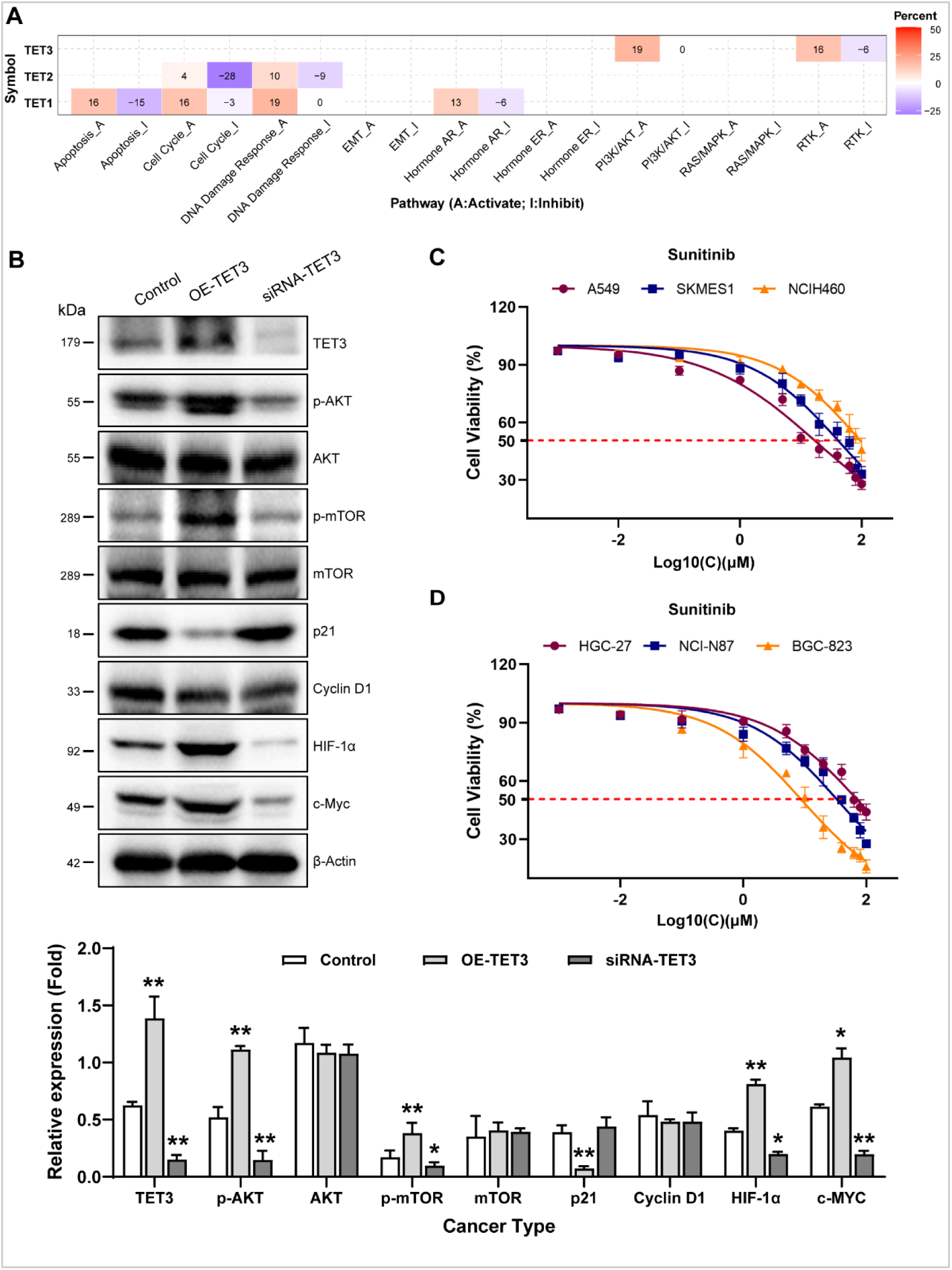
Pathway activity analysis of TET family gene in pan-cancer. (A) The combined percentage of the effect of TET genes on pathway activity. Numbers in the table represent the percentage of pathway activity on TETs. Red represents activate effect and blue represents inhibition. (B) We detected the impact of TET3 on the protein expression levels of key members of the aforementioned signaling pathways by western blot. (C-D) We stimulated different pathological subtypes of lung cancer cells and different degrees differentiation of gastric cancer cells with Sunitinib, a selective multi target tyrosine kinase inhibitor, and then tested their therapeutic sensitivity to clarify whether TET3 affects tumor behavior by regulating the RTK pathway in Fig 4A.

Considering the importance of TET3, we overexpressed and silenced TET3 in the moderately differentiated gastric cancer cell line NCI-N87, and found that overexpression of TET3 can increase the phosphorylation levels of AKT (Ser 308 and Ser 473) and mTOR (Ser2448), inhibit P21, and increase HIF1α and c-Myc protein expression, while silencing TET3 showed the opposite trend (Fig 4B).

Sunitinib is a selective multi target tyrosine kinase inhibitor. To observe whether TET3 affects tumor behavior by regulating the RTK pathway, we tested the therapeutic sensitivity of three different pathological subtypes of lung cancer cell lines (A549, SKMES1, NCIH460) to sunitinib. The results showed that A549 and SKMES1 cell lines with relatively high expression of TET3 exhibited higher sensitivity to sunitinib (Fig 4C). Similarly, we also tested the therapeutic sensitivity of gastric cancer cell lines (NCI-N87, BGC-823, HGC-27) with different degrees of differentiation to sunitinib. The results showed that cell lines with high expression of TET3 exhibited higher sensitivity to sunitinib (Fig 4D). The above results confirm that TET3 is indeed involved in the regulation of the PI3K/AKT and RTK pathways during tumor progression.

### Influence of TET gene family on survival and prognosis of pan-cancer

Next, we analyzed the correlation between the expression levels of TETs and the survival time of 33 types of tumor patients using the TCGA database. The Kaplan Meier survival curve analysis showed that high expression of TET1 in ACC, KIRP, LIHC, SARC, and STAD implies a shorter patient survival period. In LGG and THYM, low expression of TET1 implies a shorter survival period. In OV, PCPG, and UCS, high expression of TET2 implies shorter patient survival. In KIRC, low expression of TET2 implies a shorter survival period. In ACC, MESO, and PCPG, high expression of TET3 implies shorter patient survival. Low expression of TET3 in THCA implies a shorter survival period (Figure 5, Table 8)

**Figure 5:**
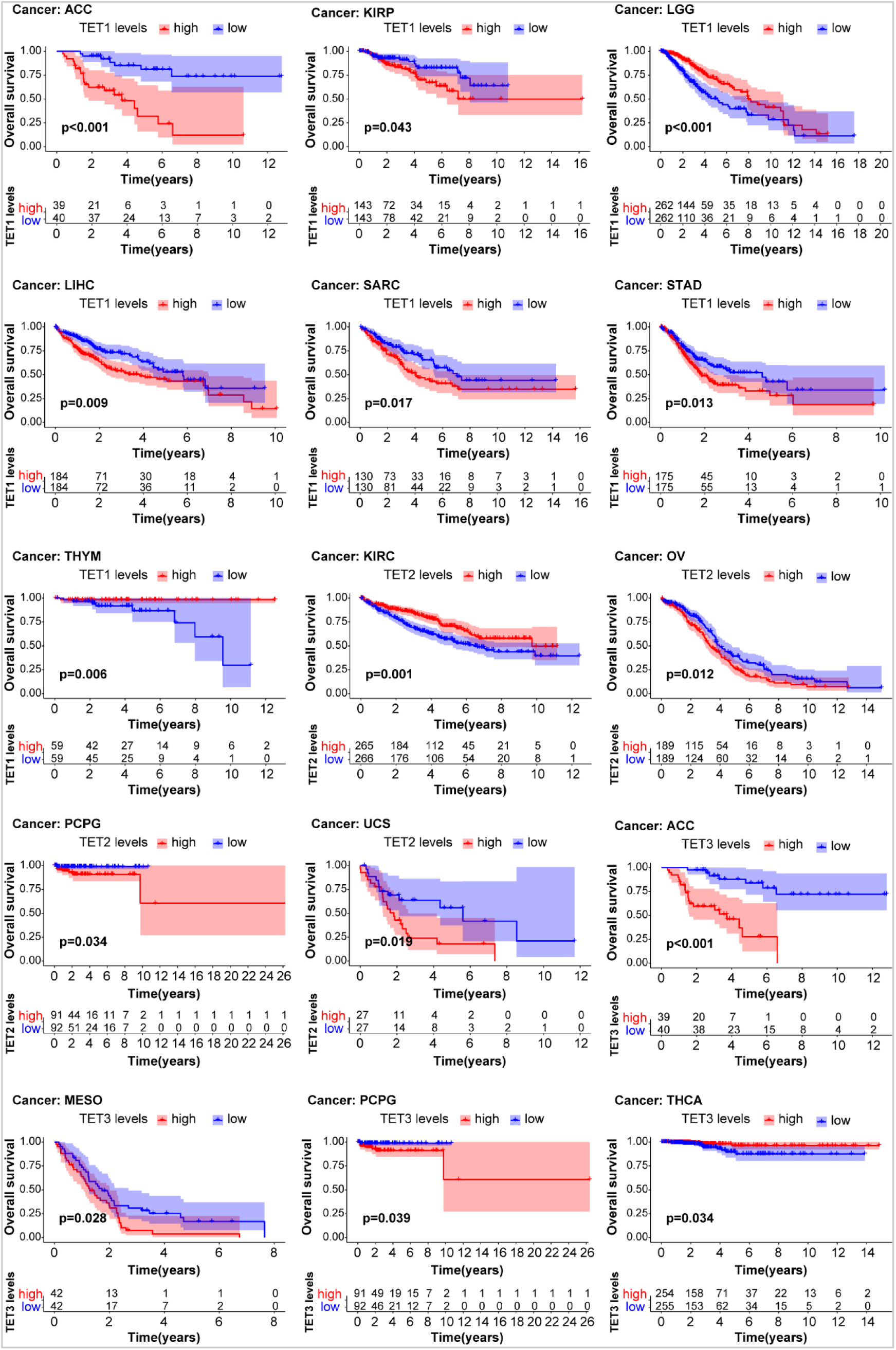
Analysis of the relationship between the expression of TET genes and the prognosis of patients with univariate KM risk proportional regression model.

On this basis, we also evaluated the relationship between the expression of TETs and the prognosis of cancer patients (Figure 6). The COX regression analysis results based on 33 types of cancer showed that the expression of TET1 in ACC, BLCA, CESC, LIHC, PCPG, and SARC indicates poor prognosis in tumor patients. The expression of TET1 in LGG, THYM, and UVM indicates a good prognosis for tumor patients. Only in KIRC, the expression of TET2 implies a good prognosis for tumor patients. The expression of TET3 in ACC alone indicates poor prognosis in tumor patients.

**Figure 6.**
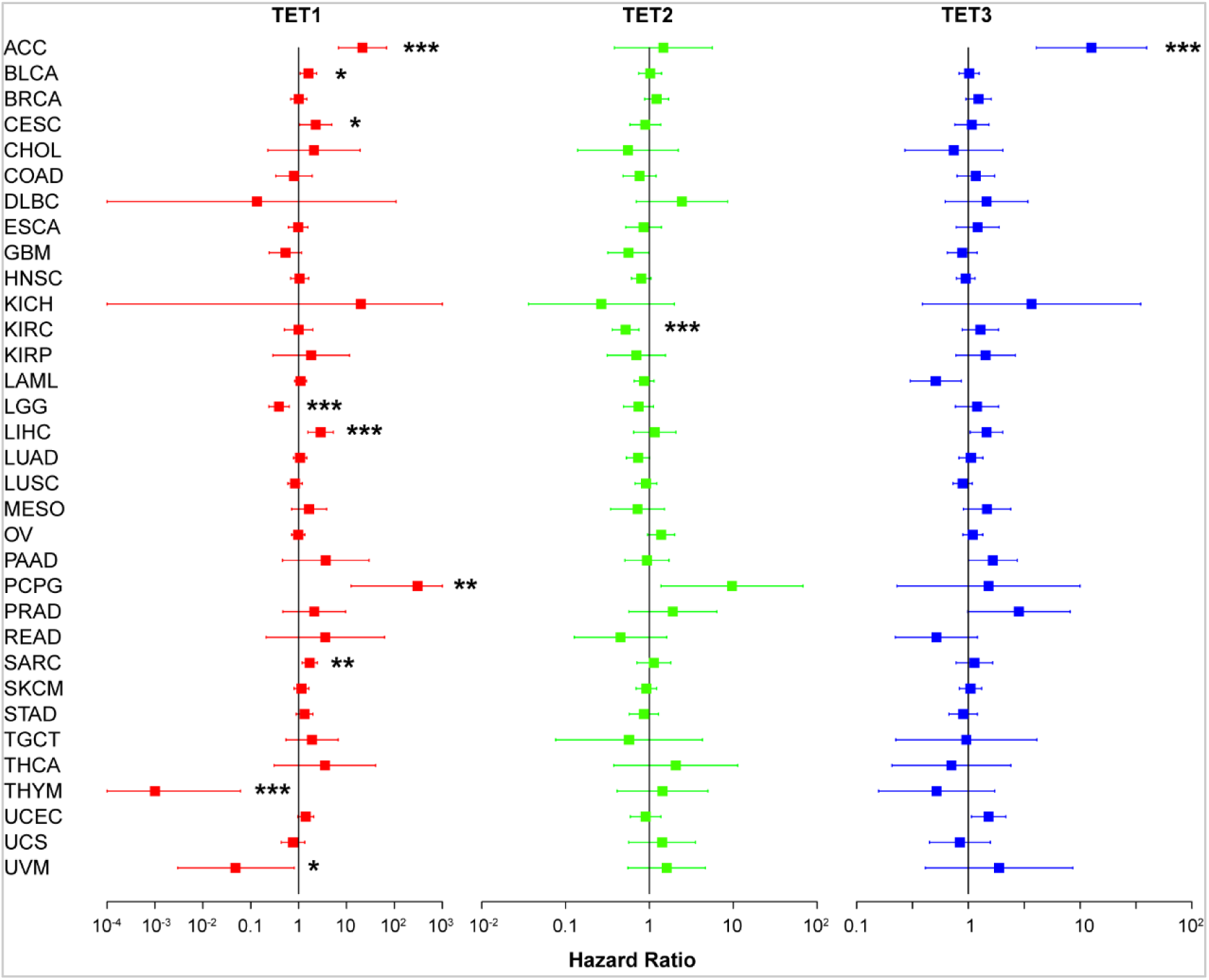
Influence of TET gene family on prognosis of pan-cancer by the COX regression analysis.

### Correlation analysis between the expression of TET family genes and immune subtypes of pan-cancer and single tumor

To clarify the relationship between TETs and immune responses, we conducted a pan-cancer analysis between TETs expression and immune subtypes based on the TACG database. As shown in the box plot of Figure 7A, the expression of TETs was significantly positively correlated with the immune subtypes of pan-cancer (*P*<0.05): among the six immune subtypes, TET1 and TET2 were found to be significantly upregulated in immune subtype C5, while their expression was stable in other immune subtypes; TET3 has varying degrees of expression with changes in immune subtypes. To clarify which immune subtypes of tumors are specifically associated with TETs, we conducted KS test correlation analysis in 8 common tumors. The results showed that except for PAAD, CHOL, and READ, the expression of TETs showed different significant correlations with the immune subtypes of other tumors (*P*<0.05), but TET1 was the lowest expressed in all immune subtypes, while TET3 was highly expressed (Fig. 7B). The expression of TETs is significantly positively correlated with various immune responses of STAD, LIHC, and LUAD. Among them, TET1 has the highest expression in the C3 subtype of STAD and is low expressed in C2. TET2 is continuously upregulated in C1-C6. TET3 has the highest expression in the C4 subtype. In LIHC, the expression of TET1 and TET3 showed a downward trend in C1-C6, but TET1 was almost not expressed in the C6 subtype. TET2 has the lowest expression in the C2 subtype. In LUAD, TET1 is expressed stably in various subtypes. TET2 has the highest expression in the C3 subtype and is downregulated to the lowest in the C4 subtype. The expression of TET3 is highest in the C1 subtype and low in C4. However, the immune subtypes of COAD and LUSC were only significantly correlated with the expression of TET1 and TET3. TET1 is highly expressed in various immune subtypes of COAD, while TET3 is the least expressed in the C3 subtype. In LUSC, TET1 has the lowest expression in C3 and C4; TET3 has the lowest expression in C6 and is stable in other subtypes. In summary, the expression of TET1 gene is very low in various tumors, and the expression of TET3 is highest in all tumors, but both are closely related to the immune subtypes of pan-cancer.

**Figure 7.**
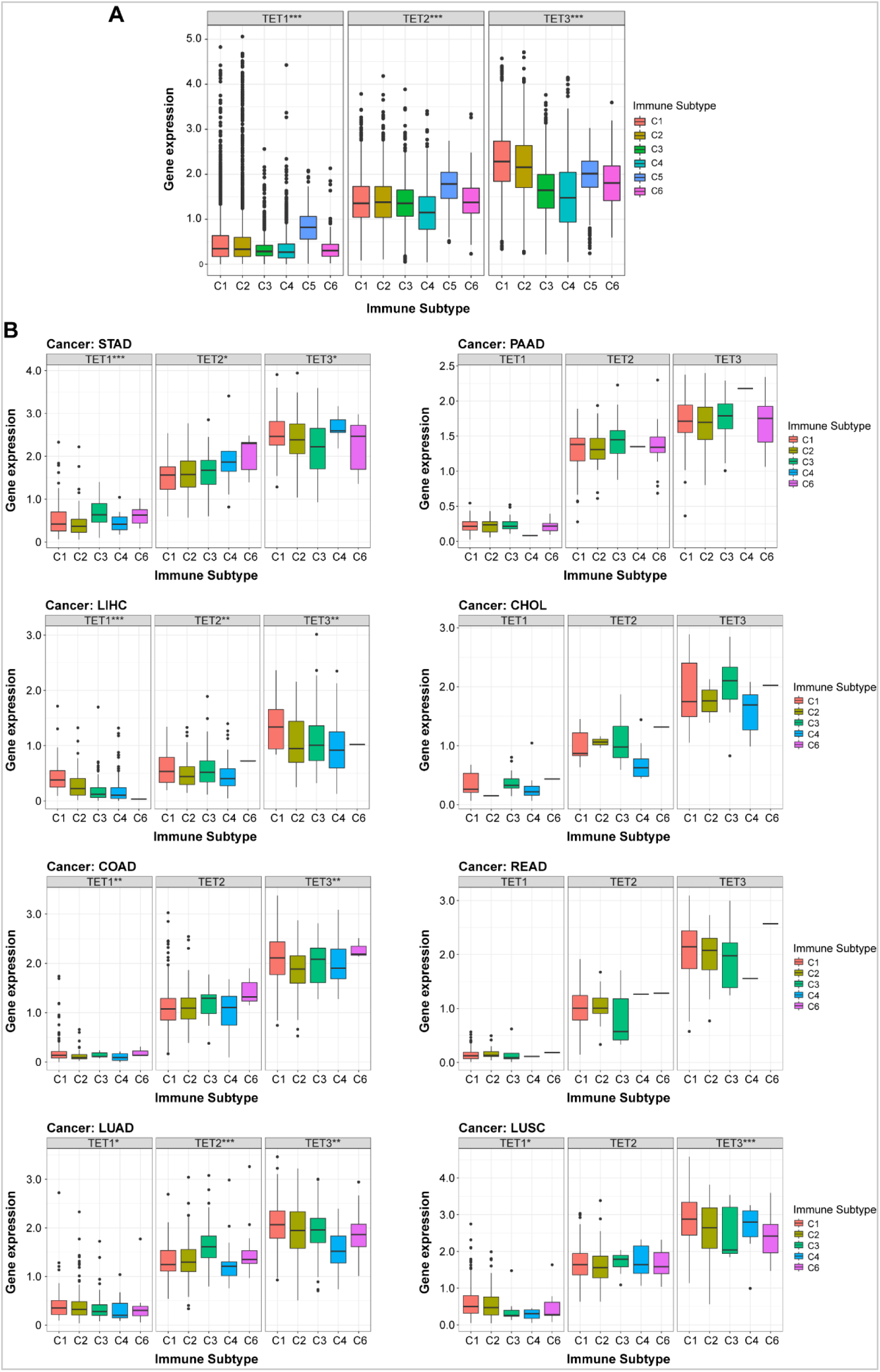
The relationship between the expression of TET gene family members and immune subtypes. (A)We analyzed the relationship between gene expression of TETs and pan cancer immune subtypes based on the TACG database. (B) Subsequently, further analysis was conducted on the correlation between the immune subtypes of 8 common tumors in clinical practice and the expression levels of TETs. Data are presented as the mean ± SD. \**P* < 0.05, \*\**P* < 0.01, \*\*\**P* < 0.001.

### Correlation between the expression of TET family genes and TME and stem cell index of pan-cancer

The effectiveness of radiotherapy, chemotherapy, and immunotherapy on tumors largely depends on TME. Here, we use the ESTIMATE algorithm to predict the TME features of 33 types of tumors by calculating Stromal Score, Immune Score, and ESTIMATE Score. From Figure 8A-C, it can be seen that TET1 has a higher Stromal Score in COAD, ESCA, KIRP, MESO, PAAD, and STAD, while it has a higher Immune Score and ESTIMATE Score in PAAD; The Stromal Score, Immune Score, and ESTIMATE Score of TET1 were lower in GBM, LGG, and TGCT. The above results suggest that increasing the expression of TET1 may improve the TME of PAAD, inhibit tumor invasion and metastasis, and increase the sensitivity of immunotherapy drugs. However, increasing the expression of TET1 may worsen the TME of GBM, LGG, and TGCT, increase their invasion and metastasis potential, and increase drug resistance.

**Figure 8.**
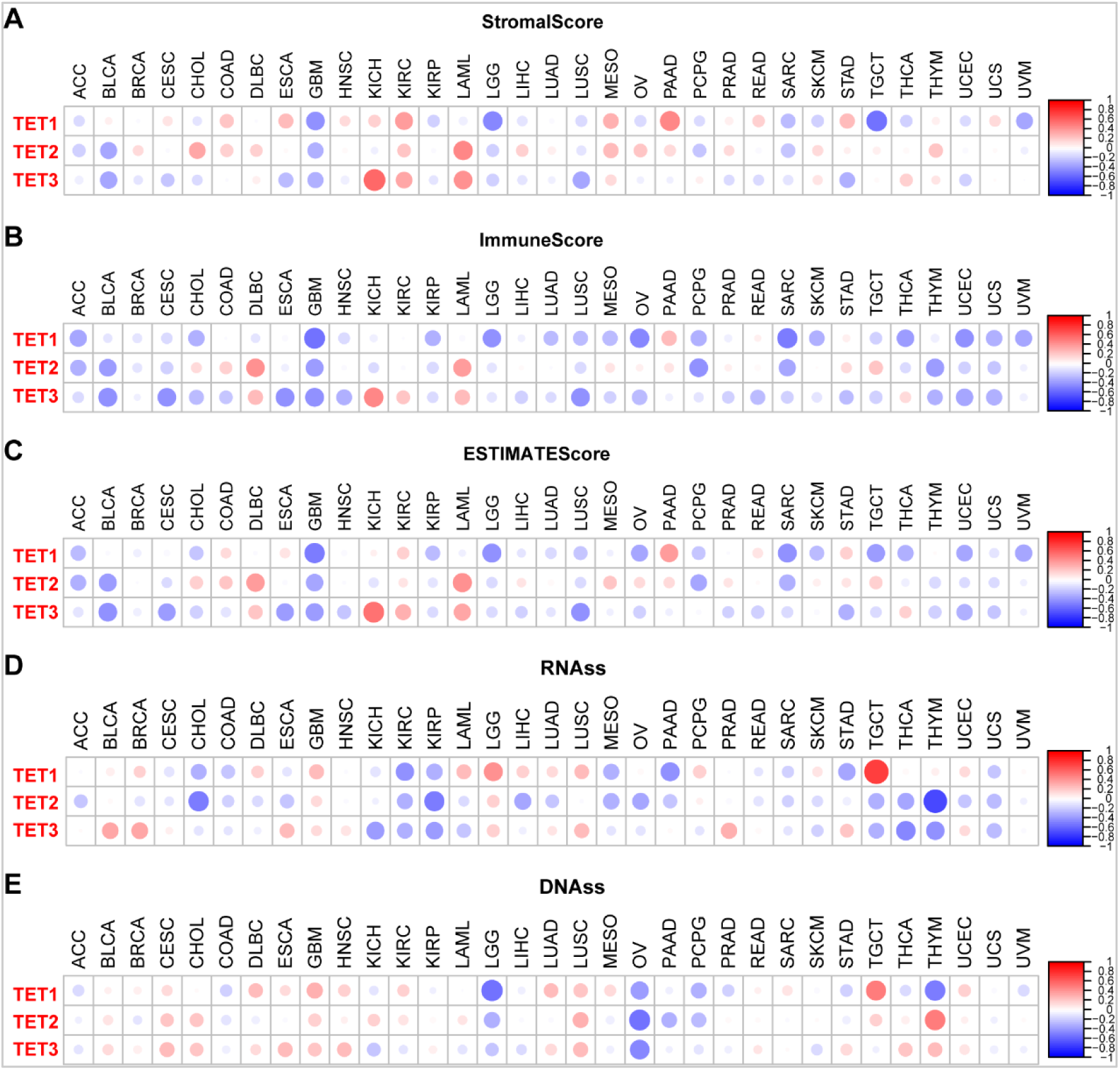
TET family members are related to tumor microenvironment of pan-cancer. (A-C) The expression of TET family genes in pan-cancer is related to tumor matrix score, immune score and tumor purity score;(D-E)The expression of TET family genes in pan-cancer is related to RNAs and DNAs.

**Figure 9.**
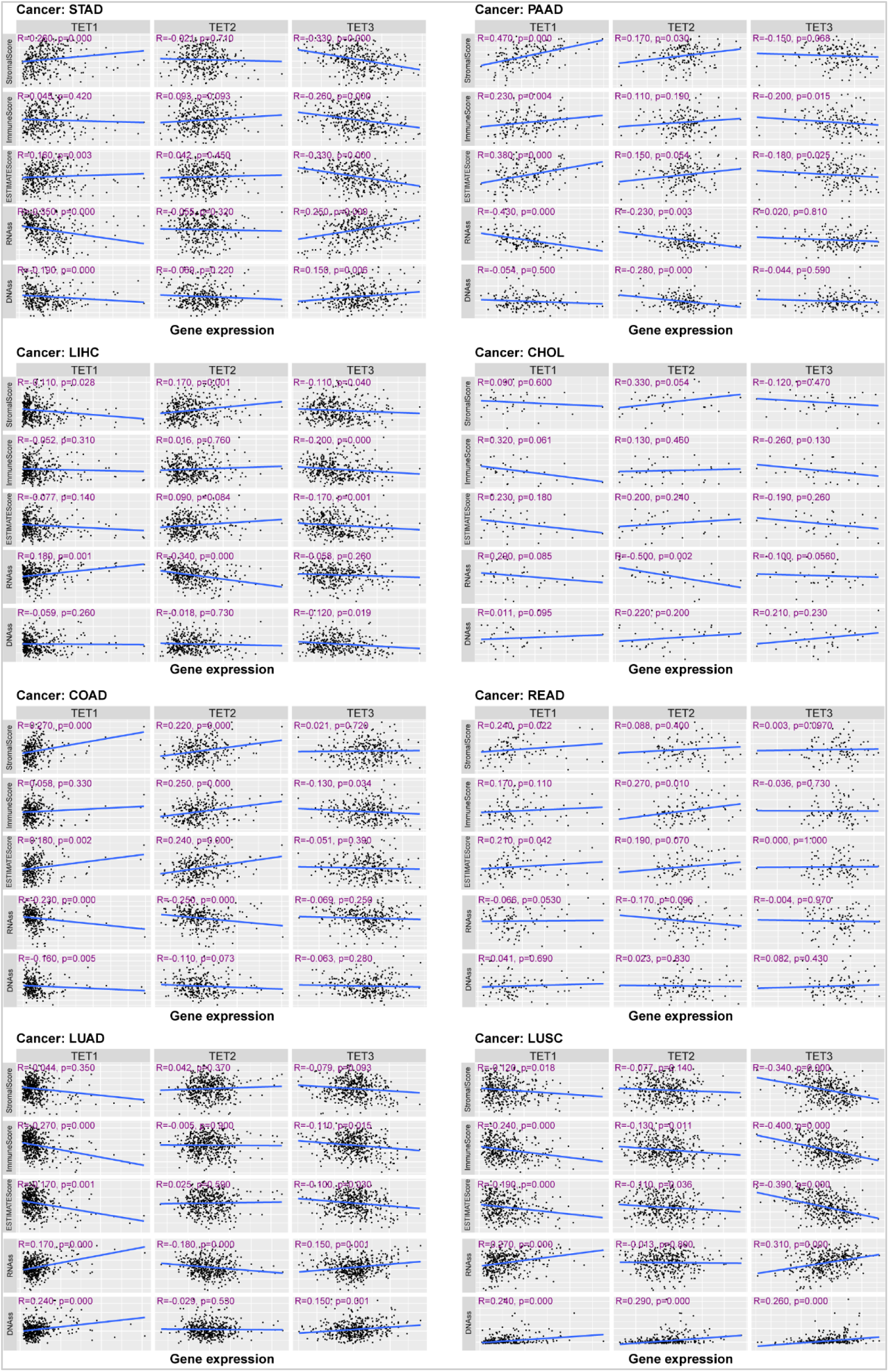
TET family members are related to stem cell index of eight common tumors in clinic.

TET2 has a higher Stromal Score in CHOL, LAML, and MESO, a higher Immune Score in DLBC and LAML, and a higher ESTIMATE Score in DLBC and LAML. It has lower Stromal Scores in BLCA and GBM, lower Immune Scores in ACC, BLCA, GBM, PCPG, SARC, THYM, and thus lower overall scores in ACC, BLCA, GBM, and PCPG. This indicates that restoring the expression of TET2 may improve the TME of LAML, inhibit tumor invasion and metastasis, and increase the sensitivity of immunotherapy drugs. However, restoring the expression of TET2 may worsen the TME of BLCA and GBM, increase their invasion and metastasis potential, and lead to drug resistance.

TET3 showed higher Stromal Score, Immune Score, and ESTIMATE Score in KICH, KIRC, and LAML; It has a lower Stromal Score in BLCA, ESCA, GBM, LUSC, and STAD, while a lower Immune Score in BLCA, CESC, ESCA, GBM, LUSC, and UCEC, resulting in a lower overall score in BLCA, CESC, ESCA, GBM, LUSC, and UCEC. It is speculated that inducing the expression of TET3 may improve the TME of KICH, KIRC, and LAML, inhibit tumor invasion and metastasis, and increase the sensitivity of immunotherapy drugs. However, inducing the expression of TET3 may worsen the TME of BLCA, CESC, ESCA, GBM, LUSC, and UCEC, increase their invasion and metastasis potential, and induce drug resistance.

In addition, the therapeutic effects of radiotherapy, chemotherapy, and immunotherapy on tumors are also closely related to the tumor stem cell index. Figure 8D-E results shows that the expression level of TET1 is positively correlated with RNAss in LGG and TGCT, while negatively correlated with KIRC, PAAD, and STAD. TET2 levels were negatively correlated with RNAss in CHOL, KIRP, and THYM. The levels of TET3 were negatively correlated in KICH, KIRP, THCA, and THYM, and positively correlated in BLCA, BRCA, and PRAD. The expression of TET1 is positively correlated with DNAss in GBM and TGCT, but negatively correlated with LGG, OV, and THYM; TET2 showed a positive correlation in TGCT and a negative correlation in OV; TET3 is negatively correlated in OV.

### Correlation between the expression of TET family genes and clinical stages of pan-cancer

These results indicate that the expression of TETs is closely related to the tumor microenvironment and tumor stemness potential. Here, we further analyzed the correlation between the expression of TETs genes and different clinical stages of 8 common types of tumors. As shown in Figure 10, there was no significant difference between the expression level of TETs genes and clinical stage of tumors in COAD and READ. The expression of TET2 and TET3 is significantly correlated with the clinical stages of LIHC, and both are highly expressed in Stage III, while downregulated in Stage IV. The expression levels of TET1 and TET3 are significantly correlated with the clinical stages of LUSC, with their expression upregulated in Stage III and downregulated in Stage IV. In addition, the TET2 level is significantly associated with the clinical staging of STAD and LUAD, with low expression in Stage II in STAD and high expression in other stage; and its levels in Stage I and IV in LUAD are higher. The expression of TET3 is also significantly correlated with the clinical characteristics of PAAD, with the lowest expression in Stage III and the highest expression in Stage IV. It can be seen that the TETs genes have a definite correlation with different clinical stages of different tumors.

**Figure 10.**
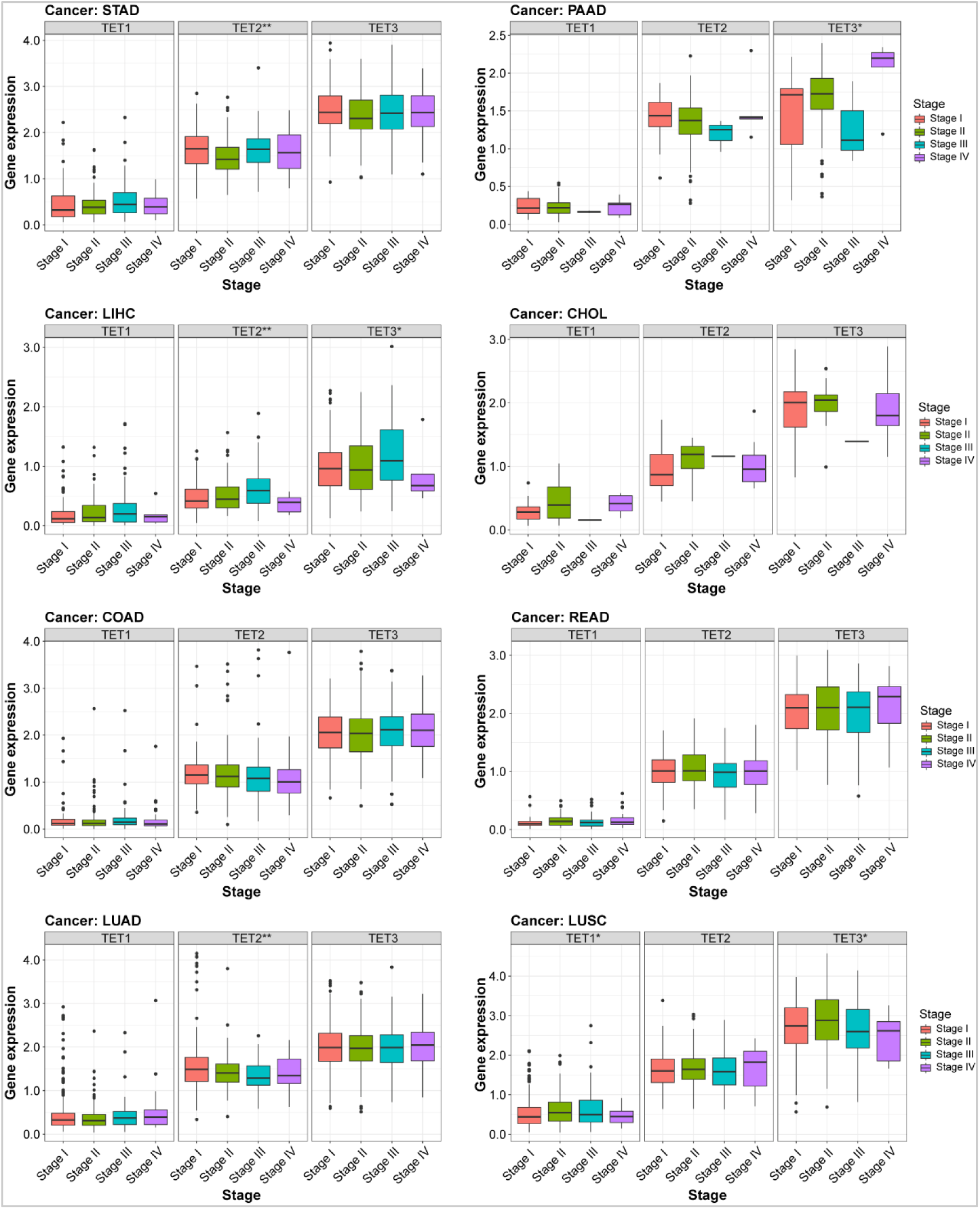
The expression of TET gene family members is closely related to the clinical characteristics of eight common tumors.

### Analysis of TET family gene levels and drug sensitivity

Using the CellMiner^TM^ database, we analyzed the potential correlation between gene expression levels and drug sensitivity of TET family members in different human tumors (Fig. 11A, Table 9), and listed the top 16 chemotherapy sensitive drugs with the highest correlation coefficient with TETs gene. Among them, the expression level of TET1 was significantly positively correlated with the drug sensitivity of Arsenal trioxide, Fenretinide, Dimethylaminophenhenolide, Daunorubicin, Homoharringtonine, Imatinib, Testolactone, Pipappererone, and Lomustine. The TET2 level is positively correlated with the drug sensitivity of Fulvestrant and Raloxifene, but significantly negatively correlated with Vemurafenib and Dabrafenib. The expression level of TET3 is significantly positively correlated with the drug sensitivity of Lapatinib, AZD-9291, and (+) - JQ1.

**Figure 11.**
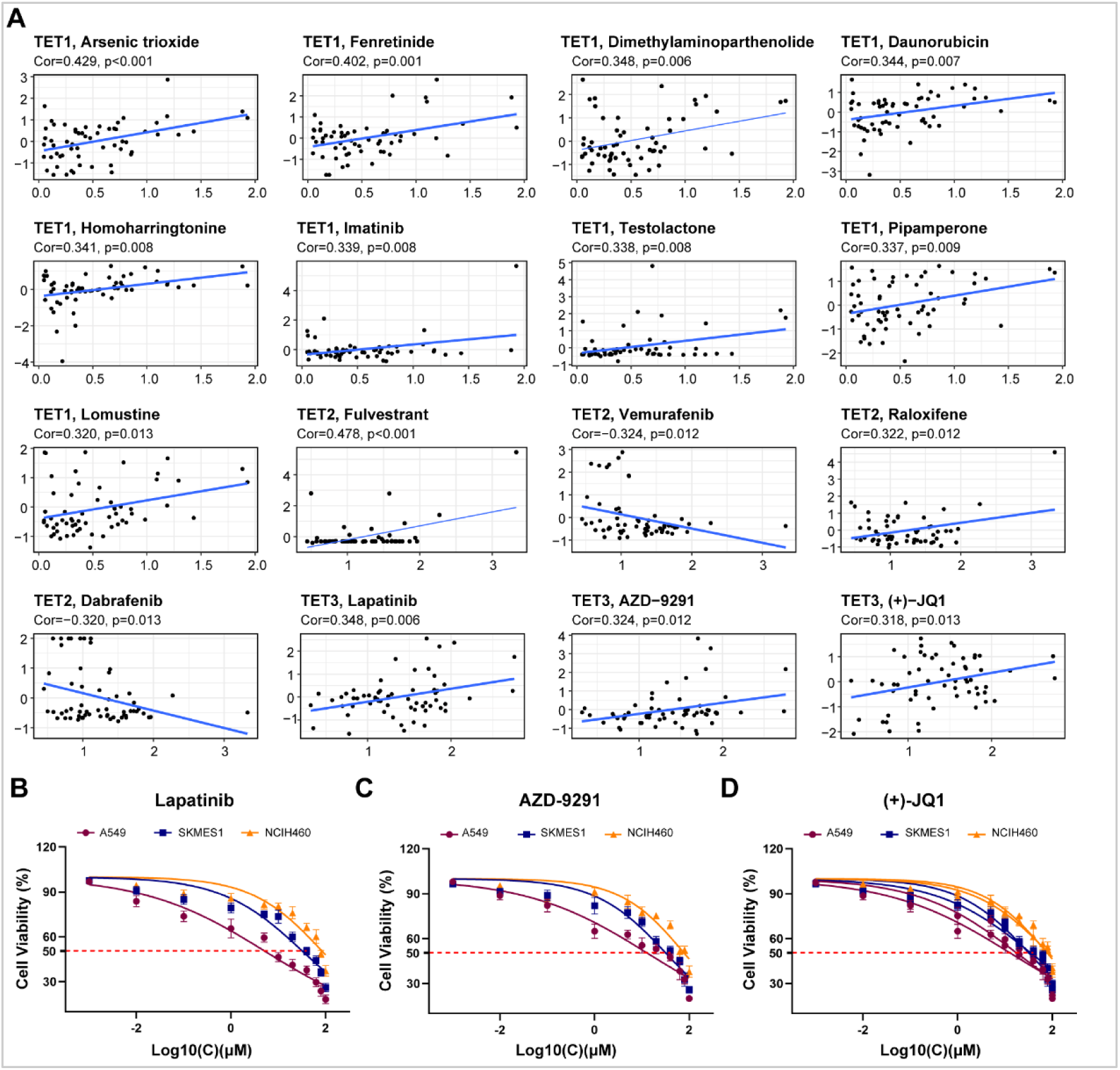
Analysis of the correlation between the expression of TET gene family members and drug sensitivity. (A) The potential correlation between gene expression levels and drug sensitivity of TET family members were analyzed based on the CellMiner^TM^ database. **(B)** We tested the therapeutic sensitivity of three different pathological subtypes of lung cancer cell lines to Lapatinib, AZD-9291, and (+) - JQ1.

We also tested the therapeutic sensitivity of three different pathological subtypes of lung cancer cell lines (A549, SKMES1, NCIH460) to Lapatinib, AZD-9291, and (+) - JQ1. The results showed that the therapeutic sensitivity of the three drugs to tumor cells was positively correlated with the expression level of TET3 (Fig 11B).

### Silencing TET3 can inhibit the malignant behavior of tumor cells

Based on the above pan cancer analysis, we speculate that TET3 is a highly potential targeted therapeutic gene. Therefore, we subsequently silenced the expression of TET3 in four tumor cell lines (BGC-823, HepG2, A549, TPC-1) (Fig 12A), and found that silencing TET3 significantly inhibited tumor cell clone formation ability (Fig 12B), migration ability (Fig 12C), and invasion ability (Fig 12D), inhibited cell proliferation cycle (Fig 12E), and increased cell apoptosis (Fig 12F). These results suggest that TET3 plays an oncogene role in various tumor progression, and targeted silencing or inhibition of the TET3 gene is expected to become a promising therapeutic option.

**Figure 12.**
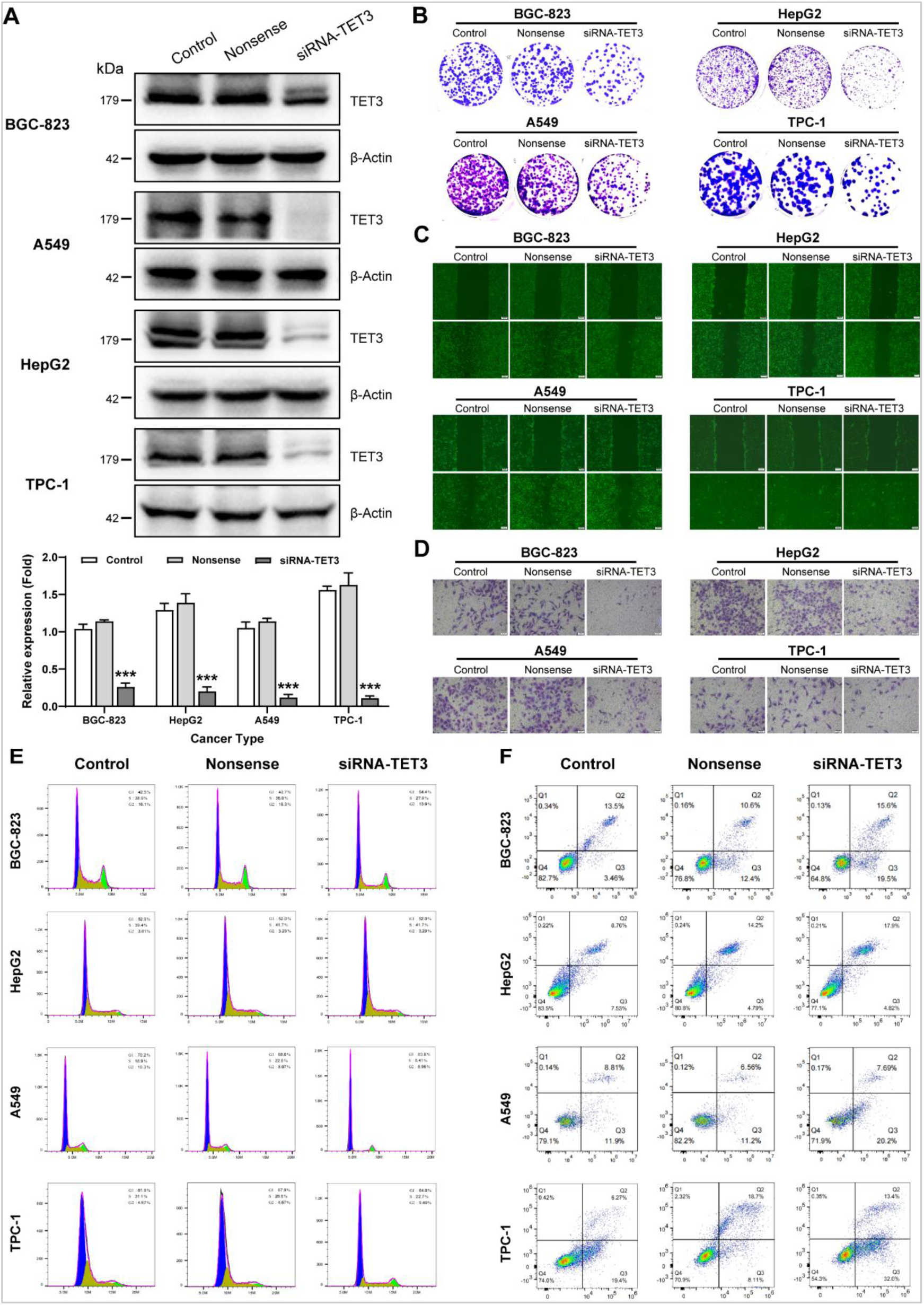
Silencing TET3 can inhibit the malignant behavior of tumor cells. (A) The expression of TET3 in four tumor cell lines (BGC-823, HepG2, A549, TPC-1) was silenced. (B-F) silencing TET3 significantly inhibited tumor cell clone formation ability (B), migration ability (C), and invasion ability (D); and also inhibited the cell proliferation cycle (E) while increased cell apoptosis (F).

## Discussion

The pattern of DNA methylation at cytosine bases in the genome is tightly linked to gene expression, and DNA methylation abnormalities are often observed in many diseases, including inflammation, hypertension, diabetes and various tumors (***Bowman & Levine, 2017; Bray, Dawlaty, Verma, & Maitra, 2021; Ismail, Ghannam, Al Outa, Frey, & Shirinian, 2020; Matuleviciute, Cunha, Johnson, & Foskolou, 2021***). Enzymes of the TET family catalyze the stepwise oxidation of 5-methylcytosine in DNA to 5-hydroxymethylcytosine and further oxidation products (***Bray et al., 2021; Delatte, Deplus, & Fuks, 2014; Joshi, Zhang, Breslin, Kini, & Zhang, 2022; Matuleviciute et al., 2021; Scott-Browne, Lio, & Rao, 2017***). These oxidized 5-methylcytosine derivatives serve as stable epigenetic modification that exert distinctive regulatory roles (***Kao, Wu, & Lee, 2016***). It is becoming increasingly obvious that dysregulation of TET protein expression or function is commonly observed in a wide range of cancers (***Kunimoto & Nakajima, 2021; F. Pan, Weeks, Yang, & Xu, 2015; Smeets et al., 2018***).

In this study, we first analyzed the transcriptome data of 33 tumors in the UCSC Xena database, and found that TET1 seems to be more specifically overexpressed in epithelioid squamous cell carcinoma (HNSC and LUSC) and hepatobiliary duct tissue (CHOL and LIHC), while it is uniformly low expressed in kidney derived tumors of various pathological subtypes (KICH, KIRC and KIRP). The TET2 gene tends to be highly expressed in adenocarcinoma (BRCA, COAD, READ and THCA). Unlike TET1 and TET2, TET3 exhibits widespread high expression in most detected tumors, except for KICH. These characteristic expression patterns may contribute to clinical diagnosis and the development of tumor targeted drugs.

Some SNPs located inside genes may directly affect protein structure or expression level, and they may represent some factors in tumor genetic mechanism (***Kuhlen et al., 2019***). In this study, we systematically analyzed the somatic cell variation landscape of TET gene family members, and found that the SNV frequency of TET family genes were 91.24%, and the main type of CNV was heterozygous amplification and deletion. Correlation analysis shows that the expression of TET2 in CHOL and ACC is positively correlated with CNV, while TET2 in LAML and TET3 in THMY are both negatively correlated with CNV. In the future, efforts should be made to screen for pathogenic CNVs and correctly interpret the clinical significance of these detected CNVs, although this work is extremely cumbersome and arduous.

In general, the degree of whole genome hypomethylation in tumor cells is closely related to disease progression, tumor size and malignancy (***Esteller & Herman, 2002***). DNA methylation detection is of great significance in judging tumor malignancy and selecting targeted drugs. The TET family genes, as the responsible genes that directly intervene in the methylation levels of many genes, their own methylation status makes gene expression regulation more complex. Although these mechanisms can significantly increase the accuracy of gene regulation, they also add research difficulties to researchers. Similar to other genes, the expression of TET family genes in pan-cancer is negatively correlated with their methylation levels, indicating that their own expression is also strictly regulated by methylation. Even so, we still found some unique events, with the TET1 gene showing high methylation in almost all analyzed cancers (except for LIHC), and the TET2 and TET3 genes showing low methylation in most tumors. Further investigation is needed in the future to determine whether this opposite expression pattern implies functional mutual compensation between TET family genes. It should be emphasized that the state of DNA methylation is not invariable in the evolution of tumors, so its clinical significance should be interpreted dynamically and carefully (***Miyamoto & Ushijima, 2003***).

Next, we analyzed the relevant signaling pathways involved in TET family genes in pan-cancer. Firstly, TET1 is mainly involved in the activation of the PI3K/AKT pathway. Xie et al. found that TET1 played an important role in promoting the proliferation of insulin-dependent endometrial cancer by up-regulating G protein-coupled estrogen receptor expression and activating PI3K/AKT signaling pathway (***Xie et al., 2017***). Huang et al. demonstrated that TET1 can control stem cell development by regulating the Wnt and PI3K-Akt signaling pathways, and TET2-deficient mice undergo a progressive reduction of spermatogonia stem cells (***Huang et al., 2020***).

Secondly, TET2 is mainly involved in cell cycle and DNA damage responses. Zhong et al. found that the 5mC oxidation is cell-cycle dependent and mainly occurs during the S and G2/M phases. TET2 depletion diminishes the observed 5hmC elevation, which suggests that the idarubicin (a topoisomerase II inhibitor) stimulation of 5hmC formation is mainly TET2-dependent (***Panigrahi et al., 2015***). Chen et al. reveals SMAD nuclear interacting protein 1 may recruits TET2 to regulate c-MYC target genes and cellular DNA damage response (***L. L. Chen et al., 2018***). Thirdly, TET3 participates in the cell cycle, apoptosis, activation and inhibition of hormone AR, and DNA damage response. Jiang and colleagues have demonstrated that DNA damage response-activated ATR kinase phosphorylates TET3 in mammalian cells and promotes DNA demethylation and 5-hydroxymethylcytosine accumulation. Moreover, TET3 catalytic activity is important for proper DNA repair and cell survival (***Jiang, Wei, Chen, Zhang, & Li, 2017***).

Then we evaluated the relationship between the expression level of TETs and the prognosis of cancer patients. Firstly, the high expression of TET1 seems to be a sign of poor prognosis in patients with solid organ derived tumors (ACC, KIRP, LIHC, and SARC), while the high expression of TET1 can be seen as a marker of good prognosis in LGG and THYM patients. Secondly, the high expression of TET2 seems to be more inclined to serve as a marker of poor prognosis in female reproductive system tumors (OV and UCS), while the high expression of TET2 implies a good prognosis in KIRC tumor patients. Thirdly, high expression of TET3 implies poor prognosis in ACC patients, but implies better prognosis in THCA tumor patients.

Next, the analysis of immune subtypes showed that TET1 was generally lower expressed in the six immune subtypes (C1-C6), but TET3 was generally higher. Other features are also displayed in this study. These results can not only characterize the heterogeneity of the immune state within tumors, but also provide reference for the selection of targeted drugs.

TME is involved in tumor survival, immune evasion, and drug resistance (***Arneth, 2019***). Therefore, we further analyzed the relationship between the expression of TETs and TME. Firstly, restoring the expression of TET1 may improve the TME of PAAD, while worsen the TME of GBM, LGG, and TGCT. Li et al. suggested that TET1 is able to suppress epithelial-mesenchymal transition and sensitize PAAD cells to 5FU and gemcitabine (***Li et al., 2020***). However, there is no research on how overexpression of TET1 affects the TME of GBM, LGG, and TGCT. Secondly, restoring the expression of TET2 may improve the TME of LAML, but worsen the TME of BLCA and GBM. Cimmino et al. found that TET2 restoration may reverse aberrant hematopoietic stem and progenitor cell self-renewal in vitro and in vivo (***Cimmino et al., 2017***). Similarly, it is not yet known how inducing TET2 expression affects the TME of BLCA and GBM. Thirdly, inducing the expression of TET3 may improve the TME of KICH, KIRC, and LAML, but worsen the TME of BLCA, CESC, ESCA, GBM, LUSC, and UCEC.

Further results based on the tumor stem cell index showed a positive correlation between the high expression of TET1 and the RNAss and DNAss of TGCT. Benešová et al. found highly increased expression of TET1 dioxygenase in most seminomas and strong TET1 staining in seminoma cells. And they propose that TET1 expression should be a potential marker of seminomas and mixed germ cell tumors (***Benešová et al., 2017***). These findings suggest that TET1 may be an important indicator for evaluating the stemness maintenance potential of tumor stem cells and chemoradiotherapy resistance in TGCT.

The analysis of the correlation between the expression of TET family genes and clinical staging of common tumors found that TET1 only showed correlation with tumor staging in LUSC, but this statistical difference is weak and seems to have no reference value for the true state. Unlike TET1, TET2 showed a significant correlation with tumor staging in STAD, LIHC, and LUAD (*P*<0.01). TET3 also showed a correlation with tumor staging in PAAD, LIHC, and LUSC (*P*<0.05). Liu et al showed that both global genomic 5hmC and 5fC contents were decreased significantly in the very early stage (stage 0) of LIHC compared with those of para-tumor tissues. Noteworthily, 5fC content continued to decrease in the late stage (BCLC staging from 0 to A) of LIHC. Moreover, the significantly positive correlations among the expression levels of TET2 in para-tumor tissues were generally attenuated or even disappeared in LIHC tumor tissues (***Liu et al., 2019***). Research by Sajadian et al clearly show that the expression and activity of TET2 and TET3 proteins but not TET1 are impaired in hepatocellular carcinoma leading to the reduction of 5hmC in LIHC (***Sajadian et al., 2015***). These conclusions are close to our analysis results.

Drug sensitivity has always been the key to individualized cancer chemotherapy. The study of drug sensitivity is crucial for achieving personalized treatment for cancer patients and promoting the development of precision medicine (***Chaudhry & Asselin, 2009***). However, due to the heterogeneity between individuals and the significant differences in drug sensitivity, the utilization efficiency of limited medical resources is low (***Panczyk, 2014***). Therefore, it is necessary to study molecules related to drug reactions to optimize drug therapy. We analyzed the relationship between TET family gene expression and drug sensitivity in this study and reported some valuable findings. According to the above results, we found that the gene expression level of TETs is related to the sensitivity of certain drugs, and the detection of TET1, TET2, TET3 expression has particular guiding significance for the clinical selection of drugs.

It is particularly emphasized that based on integrated bioinformatics analysis and molecular biology and cytology experiments, we found that TET3 may be one of the most promising targets for various tumor treatments. In the future, direct or indirect intervention measures targeting the TET3 gene itself or its pathway may produce expected therapeutic effects.

### Conclusions

Our study demonstrated the expression profile of the TET family gene in pan-cancer, and the TET family gene was related to the disease prognosis and correlated with TME and stemness score in pan-cancers. Moreover, the gene expression level of TET1, TET2, TET3 in tumor cells is related to specific drugs’ sensitivity. These findings may provide insights for further investigation of the TET family gene as potential targets in pan-cancer.

## Materials and Methods

### Data download preparation

Based on the UCSC Xena database at the University of California, Santa Cruz (https://xenabrowser.net/datapages/), RNA Seq data and clinical related data for 33 types of tumors starting with "GDC TCGA" were downloaded, including "HTSeq FPKM (n=151) GDC Hub" data under Gene expression RNAseq; The "Phenotype (n=697) GDC Hub (Clinical Traits)" and "Survival Data (n=626) GDC Hub" data under the Phenotype category. TCGA pan-cancer (PANCAN) related data, including "Immune subtype" data under Phenotype and "Stemness score (DNA methylation based) pan-cancer Atlas Hub" and "Stemness score (RNA based) pan-cancer Atlas Hub" data under Signatures, has also been downloaded. We also obtained drug sensitivity information from the CellMiner^TM^ database (https://discover.nci.mih.gov/cellminer/home.do).

### Differential analysis of gene expression

Extract, idTrans, and integrate the "HTSeq FPKM (n=151) GDC Hub" data under the Gene expression RNAseq item downloaded above using Perl software. Boxplot graph was drawn to display the expression level of TETs genes in pan-cancer tumor samples. Subsequently, samples containing at least 5 normal control samples from each type of tumor were selected to draw gene expression boxplot plots, and the "Wilcox. test" method was used to analyze the expression differences of TET family genes in different cancer types. Among them, "*", "**", and "***" respectively show p<0.05, <0.01, and <0.001. Subsequently, based on the above p-values, a heat map was further drawn using the R package "pheatmap". Finally, the correlation between genes in the TET family was analyzed using the R package "coreplot".

### Somatic mutation analysis

SNV and CNV data for 33 tumor types in the TCGA database was obtained from the Xena Functional Genomics Explorer (https://xenabrowser.net/datapages/). SNV data includes several non-silent mutations, including Missense_ Mutation, Nonsense_ Mutation, Frame_ Shift_ Del, Splice_ Site, Frame_ Shift_ Ins, In_ Frame_ Del, In_ Frame_ Ins, Translation_ Start_ Site and Multi_ Hit. Among them, the SNV mutation frequency (%) of each gene coding region is: number of mutation samples/number of cancer samples. Finally, SNV landscape maps was made by Maftools.

For CNV analysis, we divided CNV into two subtypes: homozygous and heterozygous, including amplification and deletion, respectively. Generate percentage statistics based on CNV subtypes using GISTIC processed CNV data, and calculate correlations using raw CNV data and mRNA RSEM data. Screening for genes with CNV>5% and exploring the correlation between the expression of TETs and CNV. According to the method adopted by Tyagi N et al. (***Tyagi, Roy, Vengadesan, & Gupta, 2024***), the correlation between mRNA expression and its CNV percentage sample was analyzed based on Person’s product moment correlation coefficient and t-distribution. Among them, the p-value is corrected by FDR.

### Methylation analysis

DNA methylation data is from UCSC Xena database. In differential methylation analysis, only 14 types of tumors with at least 10 paired of tumor and normal tissue were retained. Using Student’s T-test to determine methylation differences between tumor and normal samples, P-values were corrected by FDR, and FDR < 0.05 was considered significant. Subsequently, after integrating data on methylation and TETs gene expression, Spearman Correlation Coefficient was used to determine the correlation coefficient and p-value.

Combining methylation data and clinical overall survival data, the median methylation level of genes was divided into two groups and Cox regression analysis was performed to estimate the HR of gene methylation. If the Cox coefficient was>0, the survival rate of the high methylation group was worse, and the survival rate of the high methylation group was defined as high-risk, otherwise it was defined as low-risk. Meanwhile, the distribution of the two groups was compared through log rank test, and p<0.05 was considered statistically significant.

### Signal pathway activity analysis

The reverse protein array (RPPA) data from the UCSC Xena database in TCGA is used to infer marker pathway activity scores for all tumor samples in TCGA, involving 9 pathways related to cancer, including Apoptosis, Cell cycle, DNA Damage Response, Epithelial Mesenchyme Transition (EMT), Hormone androgen receptor (AR), and Hormone estrogen receptor (ER), Phosphatidylinositol-4,5-bisphosphate-3-kinase (PI3K) / protein kinase B (AKT), Rasopathes (RAS) / mitogen activated protein kinase (MAPK), Receptor Tyrosine Kinase (RTK). The pathway score is the sum of the relative protein levels of all positive regulatory components minus the protein levels of negative regulatory components in a specific pathway. Subsequently, referring to the methods of Akbani et al. and Ye et al., the pathway activity score (PAS) was estimated. When PAS (high in gene A group)>PAS (low in gene B group), gene A is considered to have an activation effect on the pathway, otherwise it has an inhibitory effect on the pathway.

### Survival and Prognostic Analysis

Based on the Survival data (n=626) GDC Hub data in the UCSC Xena database, we combined the mRNA expression data of TETs genes with corresponding clinical survival data for 33 cancer types for expression survival analysis. Kaplan Meier method and log rank test was used to evaluate the data for survival analysis (p<0.05). Firstly, we divided the tumor samples into high expression and low expression groups based on the median value of gene expression levels, and then plotted survival curves using R packets of "survivor" and "survival". Next, we apply a univariate COX risk proportional regression model to analyze the relationship between TET family gene expression and patient prognosis in pan-cancer. Finally, the "survival" and "forestplot" in the R package were used to draw forest maps.

### Immunological subtype analysis

Based on the "Immune subtype" data under the Phenotype item in the TCGA pan-cancer (PANCAN) of the UCSC Xena database, we used R packets "limma", "ggplot2", and "reshappe2" to perform immune subtype analysis on the TETs genes. We also used Kruskal (KS) test method to detect the expression differences of TETs in different immune subtypes, with p<0.05 being statistically significant.

### Analysis of tumor microenvironment and stem cell index

Based on the "Stemness score (DNA methylation based) pan-cancer Atlas Hub" and "Stemness score (RNA based) pan-cancer Atlas Hubs" data under the Signatures section of TCGA pan-cancer (PANCAN), we used R packets "estimate" and "limma" to predict the Stromal Score, Immune Score, and ESTIMATE Score of 33 types of tumors to analyze the purity of tumors. Subsequently, we conducted a correlation analysis between the gene expression level of TETs and ESTIMATE Score of 33 types of tumors using Spearman. In addition, we also conducted Spearman correlation test based on transcriptional data and stemness score (RNA expression level and DNA methylation level).

### Drug sensitivity analysis

Download drug sensitivity data based on the CellMiner^TM^ database, and use R packages "input", "limma", "ggplot2", and "ggpubr" to process and visualize drug sensitivity data. Perform correlation analysis on the data using Cor. Test. *P*<0.05 is considered to have significant drug sensitivity. Sensitivity of tumor cells to drugs was evaluated using the Cell Counting Kit-8 (CCK-8; Yeasen, Shanghai, China).

### Cell culture, plasmids, and transfections

All cell lines involved in this study (including gastric mucosal epithelial cell line GES-1, liver cell line LX2, lung epithelial cell line 16HBE, thyroid cell line Nthy-ori-3-1, poorly differentiated gastric cancer cell line BGC-823, hepatoblastoma cell line HepG2, lung adenocarcinoma cell line A549, papillary thyroid cancer cell line TPC-1, lung squamous cell carcinoma cell line SKMES1, large cell lung cancer cell line NCIH460, moderately differentiated gastric cancer cell line NCI-N87, and undifferentiated gastric cancer cell line HGC-27), were purchased from the American Type Culture Collection (ATCC; USA) and cultured according to recommended culture conditions.

TET3-targeting interfering RNAs (siRNA-TET3) and TET3 overexpression plasmid (pcDNA3.1-3*Flag-TET3) were synthesized by GenePharma Co., Ltd (Shanghai, China) and transfected at a concentration of 20 nM with Lipofectamine RNAiMax (Invitrogen, USA). The siRNA sequences were as follows: siRNA-TET3:5’-GGAAAGAGCUCCCGCGGUUTT-3’. The plasmid was constructed using Fast MultiSite Mutagenesis System Kit (FM201-01, TransGen Biotech, China) by the manual instruction. The plasmids mentioned above were transfected into cell lines with Lipofectamine 2000 reagent (Thermo Fisher Scientific, USA), and transfected cells were employed in experiments 48 h post-transfection.

### Western blot

Total protein was extracted from cells using RIPA lysis buffer and quantified with a BCA kit (Beyotime, Shanghai, China, P0010), according to the manufacturer’s operation. The denatured lysate was separated on SDS-polyacrylamide gel (SDS-PAGE) by electrophoresis and transferred to the PVDF membrane. Then membranes were blocked in 5% skimmed milk powder for 1h at room temperature, incubated with primary antibody overnight at 4℃, then incubated with secondary antibody for 1h at room temperature. Finally, Immunoreactive bands were incubated with the ECL (Thermo Fisher Scientific, 34580) for 1 min and detected on a ChemiDoc™ MP Imaging System (BioRad, Shanghai, China). Information on all antibodies used for WB are listed as follows: TET3 (Abcam, ab153724, 1:1000), β-Actin (CST, #3700, 1:1000), p-AKT (CST, #4060, 1:2000), AKT (CST, #9272, 1:1000), p-mTOR (CST, #2974, 1:1000), mTOR (CST, #2983, 1:1000), p21 (Invitrogen, MA5-14949, 1:1000), Cyclin D1 (Invitrogen, MA5-16356, 1:200), HIF-1α (CST, #3716, 1:1000), c-Myc (Invitrogen, MA1-980, 1:1000).

### Behavioral detection of tumor cells

According to the experimental requirements, the flat plate cloning experiment, wound healing assay, cell invasion assay, flow cytometry cell cycle and apoptosis detection involved in this project were all implemented according to standard methods.

### Statistics method

Each validation experiment has three replicates, and all experiments were repeated three times. All data and statistical graphs were analyzed by GraphPad software v.5.01, and expressed as mean + standard error (SEM). Student’s *t*-test was used when two independent groups are compared, while One-way ANOVA was used for comparisons among multiple groups. The data were considered statistically different when *p*<0.05 and shown as \**P* < 0.05, \*\**P* < 0.01, \*\*\**P* < 0.001, and ns: no significant.

## Data Availability

All the raw data in this article are obtained by the authors through pan-cancer analysis and validation experiments, without any reservations, and have not been reused in other articles.

## Acknowledgments

We thank the databases of TCGA, Kaplan-Meier Plotter, PrognoScan, and CellMiner™ for the availability of the data.

## Additional information

### Author contribution

Conceptualization: Xu P, Wu W; Methodology: Zhang C, Zheng J, Li Y; Software: Liu J, Xing G; Validation: Zhang S; Formal analysis: Chen H, Wang J; Data curation: Shao Z, Pan Y; Resources: Liu Y, Jiang Z; Writing-original draft: Zhang C; Writing-review & editing: Liu X, Xu P, Wu W; Project administration: Xu P, Wu W.

### Materials availability statement

Materials supporting the findings of this manuscript are available from the corresponding authors upon reasonable request. All relevant data can be found within the article or its supplementary information.

